# Disentangling infectiousness and susceptibility by age group using transmission pair data: a study of SARS-CoV-2 household transmission

**DOI:** 10.64898/2026.06.04.26354892

**Authors:** Ka Yin Leung, Fuminari Miura, Jantien A. Backer

## Abstract

**Background:** Differential contributions to transmission across age groups have been reported for many respiratory infections, including SARS-CoV-2. They are crucial for estimating the impact of age-specific interventions. Disentangling these age-dependent contributions remains challenging, as they may reflect differences in contact rates, biological susceptibility, or infectiousness.

**Aim:** We aim to jointly estimate age-specific per-contact infectiousness and susceptibility and their effect on the impact of age-specific interventions.

**Methods:** The age-specific infectiousness and susceptibility were jointly estimated in a Bayesian framework by combining contact data with transmission pair data (who-infected-whom). We applied this approach to 197,840 self-reported household transmission pairs collected in the Netherlands during the COVID-19 pandemic. Using these estimates, we projected the expected impact of school closure and work-from-home measures during the early stages of an epidemic in the absence of other interventions.

**Results:** Both infectiousness and susceptibility to SARS-CoV-2 infection were lowest in children aged 0-9 years and highest in adults over 30 years old, with 2-to 4.5-fold differences between these groups. Projected impacts of age-specific interventions indicated that school closures would reduce the reproduction number by 8% or 29% when age-specific susceptibility and infectiousness were or were not considered, respectively. Conversely, working-from-home policies would lead to reductions of 41% with and 20% without age-specific infectiousness and susceptibility.

**Conclusion:** Our method enables robust estimation of age-specific infectiousness and susceptibility. Accounting for these age heterogeneities is essential for projecting the impact of age-targeted interventions. Our approach is adaptable to other respiratory infections and can guide more tailored public health responses.

## Introduction

Respiratory infections often spread unevenly across age groups. During the COVID-19 pandemic [1], the role of children in transmitting SARS-CoV-2 was widely debated. Early in the outbreak, children were believed to be less susceptible to infection and potentially less likely to transmit the virus compared to adults [2–6]. These differences indicate that both the probability of transmitting the virus (infectiousness) and the probability of acquiring infection (susceptibility) upon an infectious contact vary by age. Recognizing age differences in susceptibility and infectiousness is crucial, as they influence the effectiveness of age-targeted non-pharmaceutical interventions (NPIs), such as school closures. Robust and early estimates for age-specific susceptibility and infectiousness are therefore essential for accurately projecting the impact of such interventions, especially for emerging infections.

Specific age groups can drive transmission for different reasons: they have more contacts, are more susceptible or more infectious. For example, if children have many contacts but are much less susceptible or infectious than adults, closing schools may have limited impact. Contact rates between different age groups can be measured in contact surveys, both under normal conditions [7] and when control measures are in place [8–10]. Age-dependent susceptibility and infectiousness on the other hand are often unknown, especially for a newly emerging infection. A key challenge is that these cannot be directly observed and need to be indirectly inferred from available data.

Two main approaches have been used to estimate how susceptibility and infectiousness vary by age. The first is to jointly estimate these parameters from household studies or contact tracing data, which contain information on confirmed cases and their (self-reported) infectors (referred to as infectees and infectors). In household studies, the infection status of each household member is observed [11–17]. In contact tracing data, only the contacts of infected individuals are traced [18–20]. A drawback of these data is the need to identify and monitor all contacts of infected individuals, which can be resource intensive, and typically, these datasets are limited in size. The second approach combines contact survey data with reported case data to estimate either the age-dependent susceptibility or infectiousness [21–25]. However, unlike household or contact tracing data, reported case data lack information on who infected whom, making it impossible to estimate both parameters independently; one must be assumed to estimate the other.

Here, we present a new method that overcomes the limitations of existing methods. Our approach uses transmission pair data, which can be collected during routine surveillance and contain information on both infector and infectee. The key strength of our method is its ability to jointly estimate age-specific susceptibility and infectiousness by integrating transmission pair data with contact survey data. We apply this approach to data from the COVID-19 pandemic in the Netherlands [26]. By combining transmission pairs in households with data on household composition from the same national setting, we derive consistent estimates of age-dependent susceptibility and infectiousness. We illustrate how these results can be used to project the expected impact of school closures and working-from-home policies.

## Methods

### Data

We analyzed COVID-19 data from the Netherlands from 1 July 2020 to 31 March 2021, a period when COVID-19 testing was widely accessible and mass vaccination had not yet begun. Unless otherwise specified, all data were stratified by age group and sex. Age was categorized into eight groups: 0-9, 10-19, …, 60-69, and 70+ years.

Test-positive case counts were obtained from Dutch registration data [27]. There were a total of 1,016,854 cases in the study period. For a subset of these cases, the most probable infector was identified, allowing construction of pairs of infectee and infector (transmission pairs) [26]. Throughout the study period, various NPIs affected community contact rates, but household structures were assumed to remain stable under these measures. Therefore, we restricted the analysis to transmission pairs in the household setting.

Transmission pair data within households were selected by considering transmission pairs for which the same postal code was recorded and in which the infectee reported a home setting or no setting where the transmission likely occurred [26]. Care homes and assisted living facilities were excluded from the household definition. This selection yielded a total of 197,861 household transmission pairs [28]. In the open data, age-sex combinations with less than 5 transmission pairs are reported as “<5” and were set to three for analysis. Validation with undisclosed data confirmed that this choice did not affect the results, as expected given the large sample size.

Household compositions (i.e., age and sex of each member in the household) were used as a proxy for the number of household contacts per day between individuals from all age-sex groups, under the assumption that each individual has daily contact with every household member. This is consistent with established approaches for projecting age-sex-specific household contacts from household composition data [29]. Data on household composition, community contacts, and seroprevalence were obtained from a nationwide serological survey (Pienter Corona), conducted during the COVID-19 pandemic in the Netherlands [10,30,31]. Household composition data were taken from all unique participants of the Pienter Corona survey. Household contacts per day by age group and sex were determined from 13,569 reported household compositions, weighted to reflect the age and sex distribution of the Dutch population [32]. As a proxy for seroprevalence, the estimated proportion infected at least once in each age group, was used. For the main analysis we used the average over rounds 3 and 4 (September 2020 and March 2021) that best coincided with our study period. For separate analyses on the periods when the wildtype and Alpha variants were dominant, we used estimates of respectively round 3 (September 2020) and round 4 (March 2021).

As validation of the method, outbreaks were simulated in both households and the community. Community contact data by age group and sex were obtained from survey round 4 (March 2021). To estimate the impact of age-targeted NPIs, we used data on daily community contacts in the absence of interventions, available from a nationwide survey conducted before the pandemic in 2016–2017 (PIENTER3) [30].

Transmission pairs and household contacts by age group and sex are shown in Fig. 1. Additional details are available in Supplemental Information Section S2.

**Figure 1:**
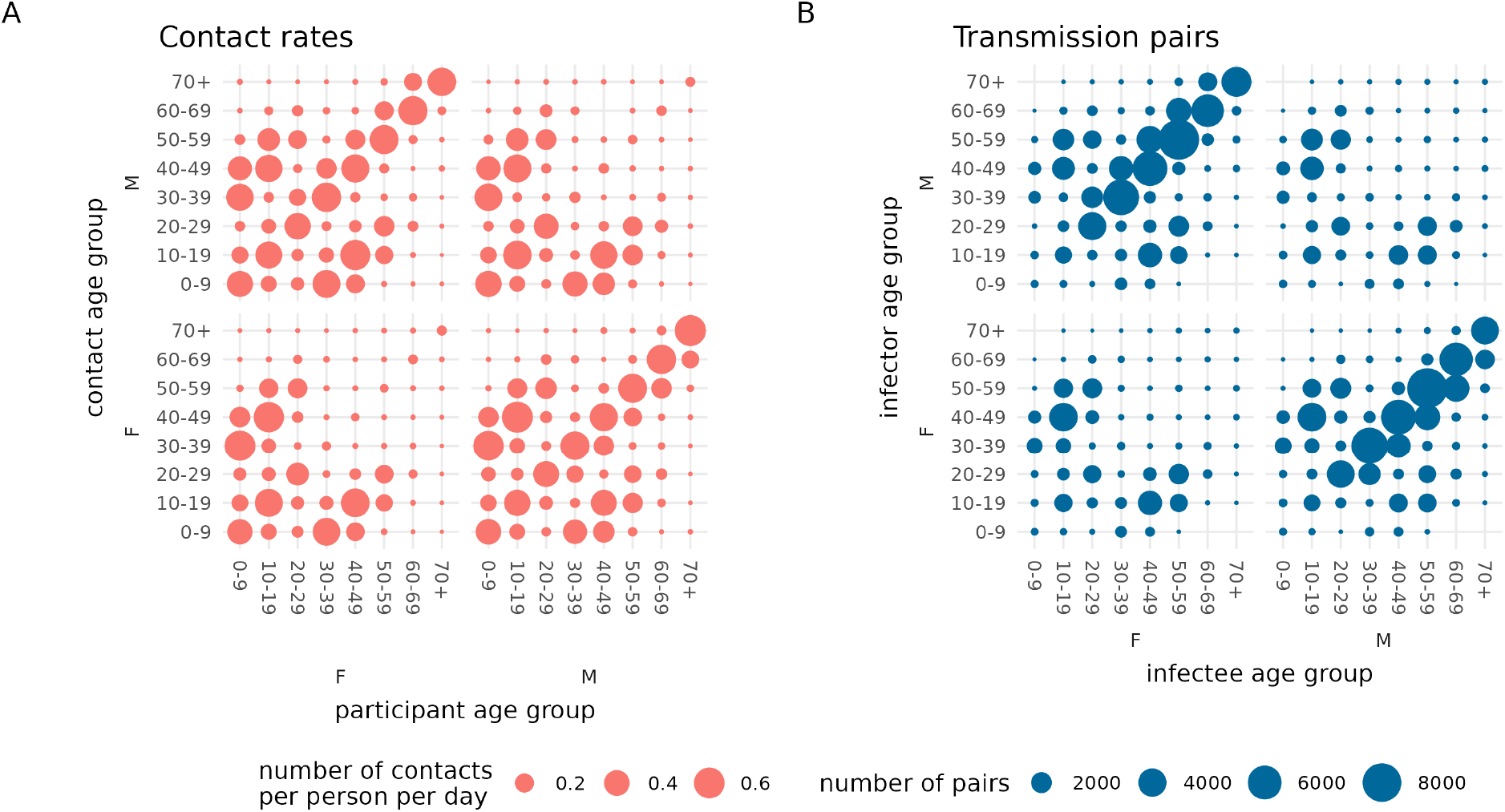
Mean number of household contacts made by individuals of one age group and sex with individuals of other age group and sex combinations (A), and household transmission pairs by age group and sex (B). F indicates female, M indicates male. Circle size indicates the contact rate (A) or the occurrence of transmission pairs (B) in each age-sex group combination.

### Estimation of age-specific susceptibility and infectiousness

Susceptibility and infectiousness were defined as the probabilities of acquiring and transmitting infection, respectively, given contact between an infectious and a susceptible individual. These probabilities may vary by age group. Age-specific susceptibility and infectiousness were expressed as percentages relative to the population average. These quantities were estimated by modelling the number of transmission pairs *P*_*ij*_, with infectees from group *i* and infectors from group *j* during the observed period. Here, *i, j* = 1, …, 2x8 correspond to all combinations of sex (*F, M*) and age groups (0-9, 10-19, …, 60-69, 70+ years).

The number of transmission pairs *P*_*ij*_ from one age group *j* to another age group *i* was modeled as being proportional (∼) to how often they interact (*c*_*i*_*j*), how susceptible the infectee is (*S*_*i*_), how infectious the infector is (*I*_*i*_), how many people can have transmitted infection 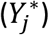, and how many people are at risk 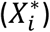:

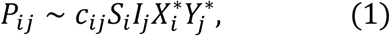

The numbers of susceptible individuals 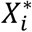 in each group *i* were assumed constant during the initial stages of the epidemic, when the majority of the population was still susceptible. 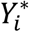 in each group *i* is the cumulative number of infectious individuals in the period in which *P*_*ij*_ was observed. The contact rate *c*_*ij*_ is the per-pair daily contact rate between an individual from group *i* and an individual from group *j*. Finally, *S*_*i*_ is the relative susceptibility of group *i* and *I*_*j*_ the relative infectiousness of group *j*.

We modeled the number of observed transmission pairs *P*_*ij*_ as a negative binomial distribution with mean 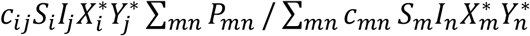 and dispersion parameter *K*, which was assigned a standard half-normal prior (note that any constant of proportionality canceled out). We applied this likelihood to the Dutch COVID-19 datasets within a Bayesian framework to estimate age-specific susceptibility and infectiousness.

We assumed that susceptibility and infectiousness were age-specific but not sex-specific, allowing estimation of age-specific susceptibility *S*_*K*_ and infectiousness *I*_*K*_, where *K* = 1, …,8 represented the eight age groups. Other terms in the likelihood were informed by the datasets described in Section 2.1. The average number of susceptible individuals 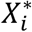 in age-sex group *i* was set equal to the number of uninfected individuals in that age group (independent of sex) averaged over the study period, informed by the serological study. The cumulative number of infected individuals 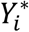 in age-sex group *i* was set equal to the cumulative number of confirmed test-positive cases in age-sex group *i* in the study period. The number of daily contacts *c*_*ij*_ between an individual in age-sex group *i* and an individual in age-sex group *j* was derived from household composition data (see Section 2.1). Using the observed number of household transmission pairs *P*_*ij*_ between age-sex groups *i* and *j*, we estimated the age-specific susceptibility *S*_*K*_ and infectiousness *I*_*K*_. The parameters *S*_*K*_ and *I*_*K*_ were assigned independent Beta(1, 7) priors (mean 1/8 = 0.125). We normalized the susceptibility and infectiousness vectors by dividing by population averages, so that the value of one corresponded to the population average. See Supplemental Information Section S1 for a detailed derivation of the model.

### Validation with simulated outbreak data

To validate our inferential method, we applied it to simulated outbreak data generated with known values of age-specific susceptibility and infectiousness. We assessed the robustness of the method to model assumptions of restricting to household contacts and household transmission pairs, and the approximation of susceptible and infectious individuals by constants 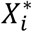 and 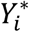, in a low-prevalence setting consistent with observed seroprevalence levels during the study period. Outbreak data were simulated using an age-structured SEIR compartmental model. In these simulations, transmission occurred through contacts in two settings: within households and in the community. Age-specific susceptibility and infectiousness were estimated from the simulated data, using only the transmission pairs and contacts in the household. These estimates were compared to the true (predefined) simulation values to assess potential bias.

In all simulations, household contact rates were as in the main analysis, derived from the household composition data. Community contact rates were informed by the reported number of contacts in the community from Pienter Corona round 4, March 2021 [10,30]. See Supplemental Information Section S3 for additional details.

### Projecting the impact of age-targeted NPIs

To assess the influence of age-specific susceptibility and infectiousness on the effectiveness of age-targeted interventions, we compared the relative change in the basic reproduction number resulting from each intervention, using both uniform and age-specific susceptibility and infectiousness. The basic reproduction number (i.e., the average number of individuals newly infected by one typical infectious individual in the initial stages of an epidemic) was calculated using the next-generation-matrix approach [33]. Two illustrative intervention scenarios were considered: 1) school closures, modeled as a 65% reduction of the number of daily community contacts among 0-19-year-olds [34], and 2) work-from-home, modeled as a 40% reduction in daily community contacts among 20-59-year-olds [35]. Reductions in the basic reproduction number are calculated assuming no other interventions and apply only to the initial stages of an epidemic. See also Supplemental Information Section S6.

For both scenarios, reductions in the basic reproduction number were projected using the age-specific susceptibility and infectiousness estimates obtained in Section “Estimation of age-specific susceptibility and infectiousness”. Total contact rates were calculated as the sum of household contacts (based on the household composition data, as in previous estimations) and community contacts (based on pre-pandemic reported community contacts from the PIENTER3 study [30]).

## Software

All analyses were conducted using R 4.4.3 [36]. Age-specific susceptibility and infectiousness were estimated with Stan using the R interface CmdStanR 2.34.1 [37]. After a warm-up period, eight chains of 2,000 iterations each were run, and convergence was assessed by confirming that all Rhat values were close to 1. The code used to reproduce the analyses and results is available at github (https://github.com/rivm-syso/transmission-pairs-susinf). An overview of data sources is provided in the Data Availability section.

## Results

As shown in Fig. 1, household contact rates and transmission pairs by age group and sex exhibited similar overall patterns. Both include a prominent diagonal reflecting interactions between individuals of the same age, and off-diagonal elements indicating contacts and transmission between children and parents. Notable age differences were observed. For example, 0-9-year-olds had many contacts but were involved in relatively few transmission events. This suggests that children are less susceptible or infectious during contact, motivating our method to estimate these age differences.

Our method is supported by simulating an outbreak where the input values for relative susceptibility and infectiousness could be accurately recovered (Supplemental Information Section S3). The average absolute difference between input and median estimated parameter was 1.7% with the largest difference in the susceptibility for 0-9-year-olds at 6.4%. Average seroprevalence across the two Pienter Corona survey rounds (March and October 2021) ranged from 3.5% in 0-9-year-olds to 15.5% in 20-29-year-olds (see Supplemental Information Table S1 for full results). We conducted simulations to validate the applicability of our inference method to the household transmission pairs, confirming that these seroprevalences were sufficiently low to allow reliable estimation of susceptibility and infectiousness across age groups (Supplemental Information Figure S4).

We applied the model to the observed data for estimation and found that younger age groups were less susceptible and less infectious (Fig. 2). Children aged 0-9 years were the least susceptible (73% less susceptible than the population average), while those aged 10-19 years were the least infectious (43% less infectious than the population average). Estimates for adults were found to be similar, with individuals aged 50-59 years the most susceptible (22% more susceptible than average) and the 60-69-year-old group the most infectious (25% more infectious than average).

**Figure 2:**
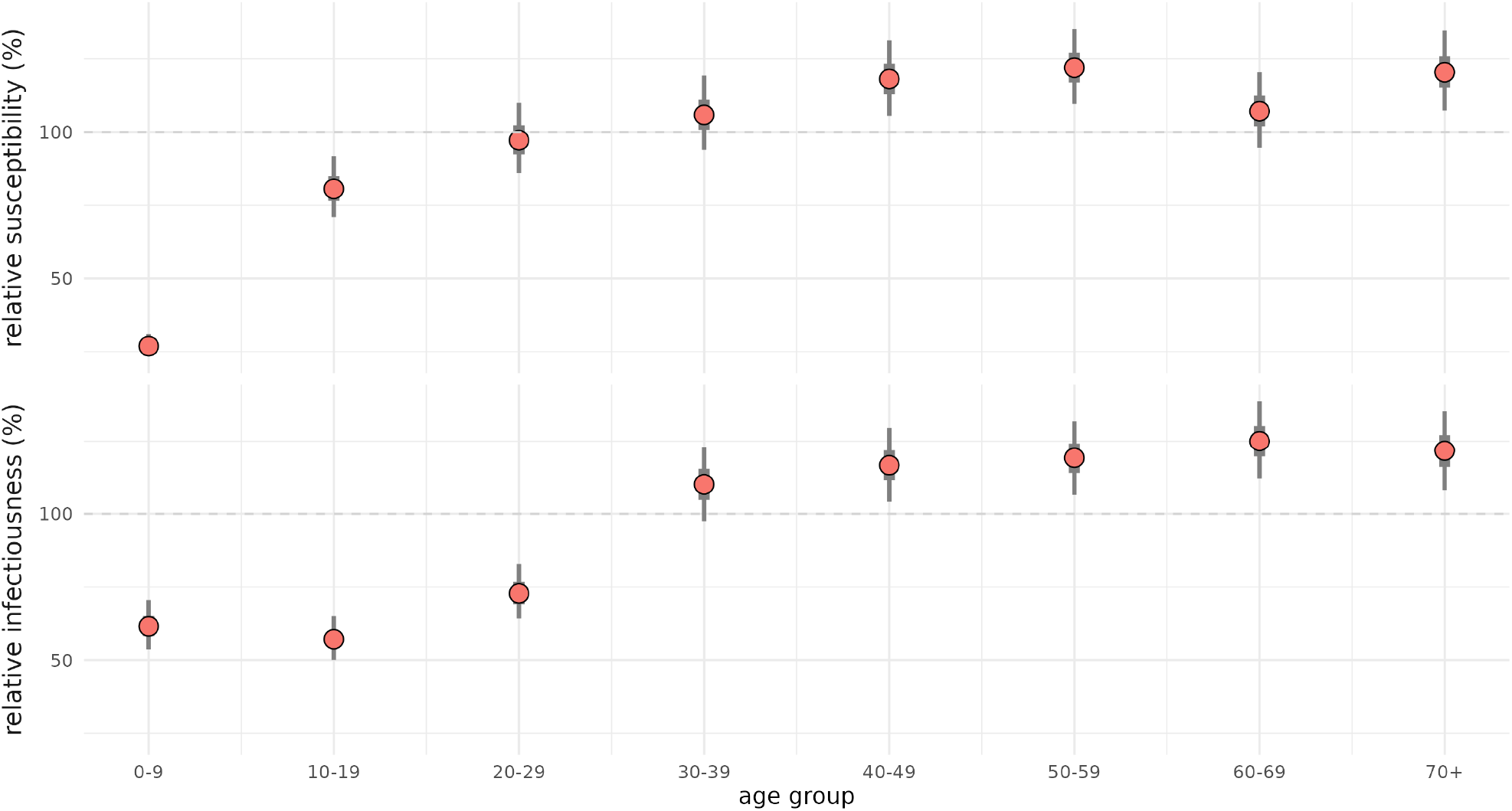
Estimates of age-specific susceptibility and infectiousness by age group, relative to the population average (in %). Points represent posterior medians; error bars denote 95% credible intervals. The dashed line at 100% indicates the population average.

The estimated age-specific susceptibility and infectiousness (Fig. 2) substantially affected projections of the effectiveness of age-specific measures. First, for a school closure scenario where 0-19-year-olds were targeted, the reduction in the reproduction number was 8% when taking the estimated relative susceptibility and infectiousness into account. In contrast, the reduction would be much larger at 29% if one would neglect the age differences in susceptibility and infectiousness. Second, for a work-from-home scenario targeting 20-59-year-olds, we found the reverse effect in the reduction in the reproduction number of 41% with the estimated relative susceptibility and infectiousness and 20% under the assumption of uniform susceptibility and infectiousness.

## Discussion

In this study, we developed a method to jointly estimate relative age-specific susceptibility and infectiousness during the early stages of an epidemic, using a combination of transmission pair and contact data. When applied to the COVID-19 epidemic in the Netherlands, our method revealed that children were both less susceptible and less infectious than adults. Additionally, our analysis showed that age-specific estimates of susceptibility and infectiousness have important implications for evaluating interventions such as school closures and work-from-home policies. Our scenarios indicated that both underestimation and overestimation of intervention effectiveness can occur if age-specific susceptibility and infectiousness are not properly accounted for.

The main strength of our study lies in its methodology, which offers several advantages over existing approaches. First, our study integrated centralized Dutch case registry data with additional data sources collected under comparable conditions and within the same time frame. This systematic approach ensured consistent observations within a single study population, without needing to rely on literature-based estimates. Second, another key strength lies in the derivation of estimators for age-specific susceptibility and infectiousness, allowing adjustment for potential age-related differences in ascertainment and asymptomatic infection (Supplemental Information Section S4). We demonstrated that our method yields unbiased estimates of age-specific infectiousness, while estimates of susceptibility may reflect age-specific variation in case detection and symptom presentation. Third, our method is general in that it can be applied to respiratory infections other than SARS-CoV-2. Finally, by focusing the analysis on household transmission pairs and household contact rates, we avoided the effects of (changing) NPIs on community contact rates.

There are several limitations to our method. First, we described the transmission within households as an age-structured compartmental model, which does not necessarily account for further heterogeneity in contact rates or individuals’ roles within a household (e.g., mothers may have higher contact rates with children [7]). However, comparing the number of household transmission pairs to household contact rates supports the choice of transmission model as a reasonable approximation of the epidemic (Fig. 1). Second, we did not explicitly account for possible age differences in asymptomatic infections or ascertainment rates for SARS-CoV-2, both of which have been shown to be age-specific [38,39]. Asymptomatic individuals are generally thought to be less infectious than symptomatic individuals [38], and differences in symptom presentation may be associated with age-related variation in ascertainment rates [39]. Susceptibility estimates cannot be disentangled from ascertainment and/or asymptomatic infections unless estimates for the latter two are available from independent data sources. Therefore, susceptibility estimates should be interpreted in light of these limitations.

Our results are consistent with previous epidemiological studies, which estimated that children are 33% to 50% less susceptible and 31% to 63% less infectious than adults [3,5,6,14,15,23]. While our primary analysis focused on the entire study period (1 July 2020 to 31 March 2021), other studies have suggested that differences in susceptibility and infectiousness between children and adults were smaller during the Alpha period compared to the wild-type period [40–43]. Our results for variant-specific periods were consistent with these findings ((Supplemental Information Section S5). Additional support comes from studies showing that children are less infectious than average due to lower virus shedding [44], and less susceptible due to a stronger innate immune response [45]. These consistent results provide further validation of our method.

Our study highlights the practical value of transmission pair data for outbreak response. Our method provides a way to leverage information from transmission pair data, which can be collected during routine surveillance. Existing methods frequently rely on data from household or contact tracing studies. Given the limitations of such approaches, if transmission pair data from routine surveillance is available, then our method allows for joint estimation of age-specific infectiousness and susceptibility estimates in a straightforward way early in an epidemic. Moreover, transmission pair data can also be used to infer other important age-specific quantities, such as serial intervals [26]. Such estimates can aid practical infection control, e.g., in relation to the generation time estimates that are crucial for estimating the speed required for effective contact tracing and quarantine [46].

In conclusion, we developed a robust method to jointly estimate age-specific susceptibility and infectiousness early in an outbreak. We demonstrated that the effectiveness of age-specific interventions against SARS-CoV-2 heavily depends on accounting for age-specific susceptibility and infectiousness. Our method is applicable to outbreaks of respiratory infections and can guide more tailored public health responses by providing more reliable projections of intervention effectiveness.

## Data Availability

All data produced are available online at https://zenodo.org/records/20327154

https://github.com/rivm-syso/transmission-pairs-susinf

## Data and code availability

Data is publicly available:

- Contact data available at [Zenodo](https://doi.org/10.5281/zenodo.10370353) [30].
- COVID-19 case data available at [RIVM open data](https://data.rivm.nl/meta/srv/dut/catalog.search#/metadata/2c4357c8-76e4-4662-9574-1deb8a73f724) [27].
- Seroprevalence data available at [GitHub](https://github.com/rivm-syso/effectiveness_NPIs) [31].
- Transmission pair data available at [Zenodo](https://doi.org/10.5281/zenodo.18269558) [28].
- Population age and sex distribution available at [CBS](https://opendata.cbs.nl/statline/#/CBS/nl/dataset/83482NED/table?ts=1747299215541) [32].

All code to reproduce the results, including downloads of the data, are available at GitHub (https://github.com/rivm-syso/transmission-pairs-susinf).

## Acknowledgements

We thank W. Speé, D. Klinkenberg, and J. Wallinga for useful feedback on the manuscript. We thank W. Speé for reviewing the codebase.

