## Supplementary information for "Disentangling infectiousness and susceptibility by age group using transmission pair data: a study of SARS-CoV-2 household transmission"

#### 1 Model derivation

Below, we provide a detailed derivation for the approximation used in the main text for the number of observed transmission pairs:

$$P_{ij} \sim c_{ij} S_i I_j X_i^* Y_j^*. \qquad (1)$$

We start by assuming an age-structured SEIR compartmental model [1]. At any given time $\tau$ during the epidemic, the number of transmission pairs per unit time with infectees from age group $i$ and infectors from age group $j$, $\pi_{ij}(\tau)$, is proportional to $c_{ij} S_i I_j X_i(\tau) Y_j(\tau)$. Here, $c_{ij}$ denotes the contact rate between an individual from group $i$ and an individual from group $j$, independent of $\tau$, $S_i$ denotes the relative susceptibility of an individual in group $i$, $I_j$ the relative infectiousness of an individual in group $j$, and $X_i(\tau)$ and $Y_j(\tau)$ the number of susceptibles in group $i$ and infectives in group $j$ at time $\tau$, respectively. In a given time period $[T_1, T_2]$, the cumulative number of transmission pairs $P_{ij}$ is therefore

$$P_{ij} = \int_{T_1}^{T_2} \pi_{ij}(\tau) d\tau \sim c_{ij} S_i I_j \int_{T_1}^{T_2} X_i(\tau) Y_j(\tau) d\tau.$$

In the study period $[T_1, T_2]$ = [1 July 2020, 31 March 2021], we approximate the number of susceptibles $X_i(\tau)$ by $X_i^*$, the number of uninfected individuals in group $i$ averaged over $[T_1, T_2]$. This approximation is reasonable early in the epidemic when the majority of the population is still susceptible, as we will show in our simulations (Section S3). With this, the equation simplifies to

$$P_{ij} \sim c_{ij} S_i I_j X_i^* \int_{T_1}^{T_2} Y_j(\tau) d\tau.$$

Next, we approximate the integral of number of infectious individuals over $[T_1, T_2]$ as being proportional to $Y_j^*$, the cumulative number of cases of group $j$ during the study period. Any constant factors are absorbed in the proportionality constant.

By applying these approximations, our model to estimate the age-specific (relative) susceptibility and infectiousness reduces to Eq. (1) above. Normalizing left- and right hand side of the model Eq. (1), the number of observed transmission pairs is modelled as a negative binomial likelihood with mean

$$c_{ij} S_i I_j X_i^* Y_j^* \sum_{mn} P_{mn} \,/\, \sum_{mn} c_{mn}\, S_m I_n X_m^* Y_n^*$$

and dispersion parameter $k$ that follows a standard half-normal distribution. Relative susceptibility and infectiousness $(S_i)$ and $(I_j)$ are assumed to be vectors that sum to 1.

#### 2 Data

Our data on transmission pairs, contacts, and cases are all stratified by age group and sex. Therefore, we use the same age group and sex stratification for the model. However, we assume that there is no biological difference in infectiousness and susceptibility by sex. Therefore, we estimate these quantities for both sexes combined, e.g. $S_{M,0-9} = S_{F,0-9}$. This means we estimate age-specific susceptibility $(S_k)_{k=1}^{8}$ and infectiousness $(I_k)_{k=1}^{8}$, for each of the eight age groups ($k = 1, \ldots, 8$).

We restrict the analysis to transmissions and contacts occurring within households. This allows us to ignore the effects of NPIs on transmissions and contacts outside of the household. Furthermore, we assume that, within a household, each individual has daily contact with every other member of the household. In reality, transmissions occur both at home and in the community, but we demonstrate (via simulation) that restricting to the household setting is a reasonable choice (Section S3).

We define a household as a group of individuals living at the same address, excluding care homes, assisted living and other institutional living arrangements. This filtering ensures that our transmission pair data reflect only transmission that occured within private households.

For each age group, we calculate the average cumulative infected fraction using data from two national survey rounds: round 3 (September 2020) and round 4 (March 2021) [2]. For completeness, the average cumulative infected fraction in each age group is shown below. In the analysis, we assumed that these proportions were the same for men and women. The cumulative infected fractions are sufficiently small to apply our method to.

Table 1: Cumulative infected fraction by age group, averaged over rounds 3 (September 2020) and 4 (March 2021) of the Pienter Corona study, used in the main analysis.

| age group | cumulative infected fraction |
|---|---:|
| 0-9 | 0.03353828 |
| 10-19 | 0.07865944 |
| 20-29 | 0.15472276 |
| 30-39 | 0.08813879 |
| 40-49 | 0.08708392 |
| 50-59 | 0.08900875 |
| 60-69 | 0.06950623 |
| 70+ | 0.07155027 |

#### 3 Simulation study to support the estimation method

To validate our inferential method, we applied it to simulated outbreak data in which the true age-specific susceptibility and infectiousness values are known. The method accurately recovers the true parameter values under realistic conditions, including low-prevalence settings consistent with observed seroprevalence levels from the two Pienter Corona survey rounds (Table S1). These results confirm the applicability of our approach to empirical outbreak data. The details of the simulation study are described below.

We used an age-structured SEIR compartmental model to simulate the transmission dynamics in the population. In this model, individuals are classified as Susceptible, Exposed (latent - infected but not yet infectious), Infectious, or Recovered (and immune). Because we focus only on the early stage of the epidemic, no waning of immunity is taken into account. In the simulations, transmission can occur through contacts in one of two settings: within households and in the community (outside the household).

The simulation uses the following input data:

- Contacts:
    - Household contacts: Contacts in the household setting are made between the sex-age groups according to the household contact matrix $c_{ij}^{H}$ (see Section Data in the main text).
    - Community contacts: The community contact matrix $c_{ij}^{NH}$ is derived from the same Pienter Corona data source (round 4, March 2021) [3,4]. Because this matrix does not distinguish between males and females, we assume that community contacts are independent of sex.
    - Total contacts: For each sex-age group pair, the total contact rate $c_{ij}$ is the sum $c_{ij} = c_{ij}^{H} + c_{ij}^{NH}$. Contacts by age group, sex and setting are shown in Fig. S1.
- Population age-group-sex distribution: the distribution for the Dutch 2021 population of 17475415 persons is obtained from Statistics Netherlands [5].
- Relative infectiousness and susceptibility: For the simulation we specify the “true” (pre-defined) values of relative infectiousness and susceptibility for each age group (see Tab. S2).

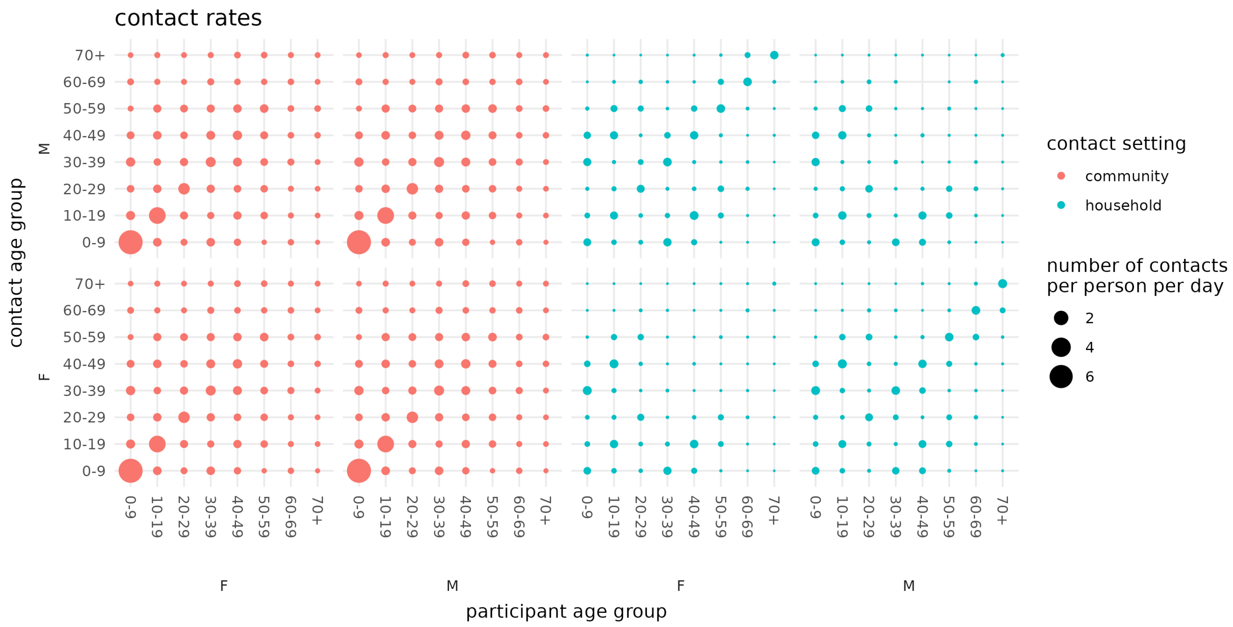


Figure 1: Household and community contact matrices. 'F' indicates female and 'M' indicates male. The household contact matrix is based on ten survey rounds of the national Pienter Corona study (April 2020–May 2023); the community contact matrix is based on round 4 (March 2021). See also the Data section in the main text and references [2–4].

Using these inputs, we simulate an epidemic. We set transmission parameters such that the following hold:

- Mean latent period equal to 2 days,
- Mean infectious period equal to 2 days
- Basic reproduction number equal to 2

The course of the simulated epidemic is presented in Fig. S2.

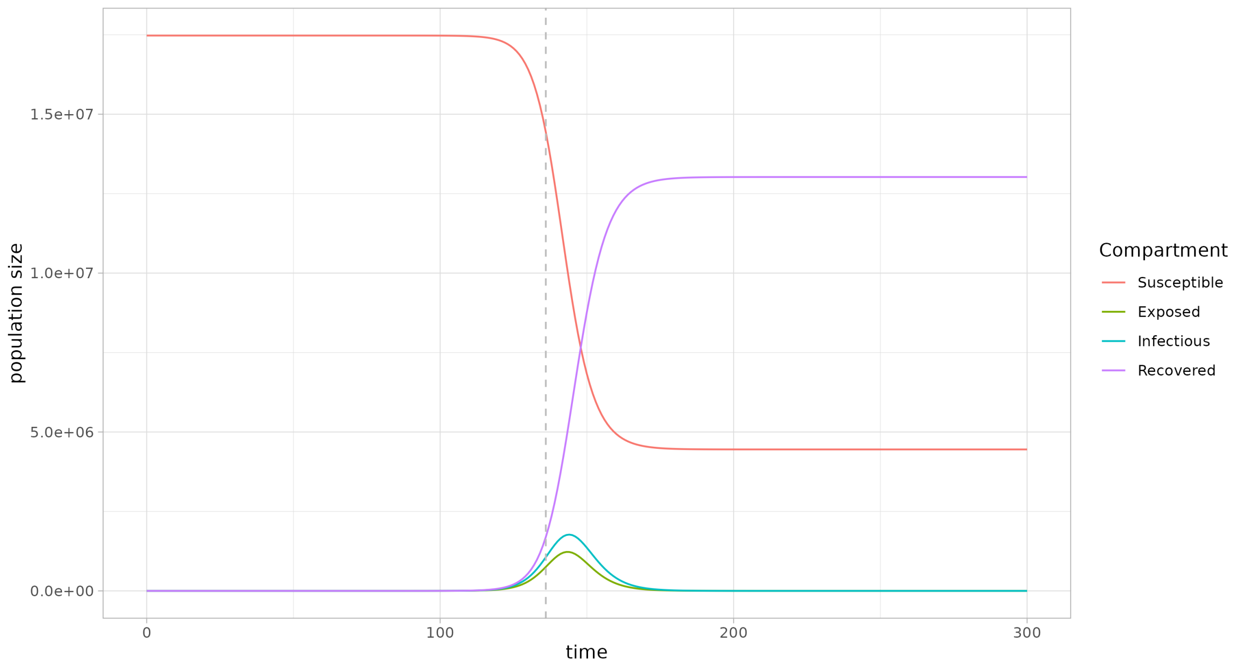


Figure 2: Course of the simulated epidemic. The dotted vertical line indicates the point when 20% of the population has been infected.

To estimate age-specific susceptibility and infectiousness, we extract data from the simulation as follows:

- The early stages of the epidemic is defined by the period that the total percentage of susceptible population that has been infected is less than 20%.
- For estimation, we use only transmission pairs resulting from household transmission $P_{ij}^{H}$ (i.e., community transmission pairs are not used); see Fig. S3.
- The total number of recovered cases (infected both through household and community transmission) during this period corresponds to $Y_j^*$.
- The recovered cases by age group are averaged over the beginning of the epidemic and used as proxy for the average never infected population $X_i^*$, $X_i^* = 1 - Y_i^*/2$.

Note that, even though only household contacts and household transmission pairs are used, community transmission effects are still incorporated in the quantities $X_i^*$ and $Y_i^*$ in the estimation.

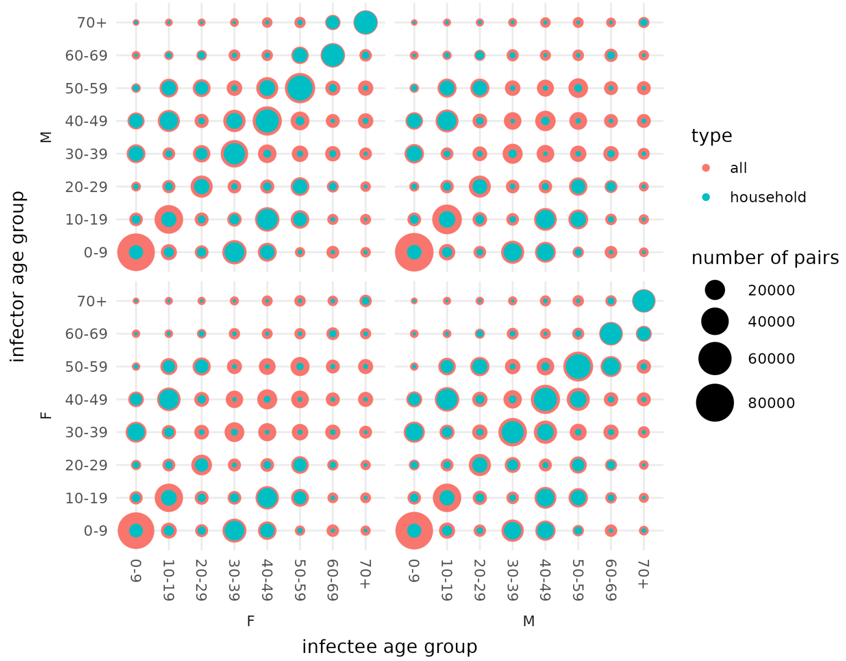


Figure 3: Simulated transmission pairs during the early epidemic period (infection attack rate <20%): comparison of household transmission pairs to total transmission pairs (including community transmission pairs.

Using these observed quantities and the household contact rates $c_{ij}^{H}$, we estimate age-specific susceptibility and infectiousness from Eq. (1).

Figure S4 and Table S2 demonstrate that our estimation method successfully recovers the true values of age-specific susceptibility and infectiousness from the simulated data.

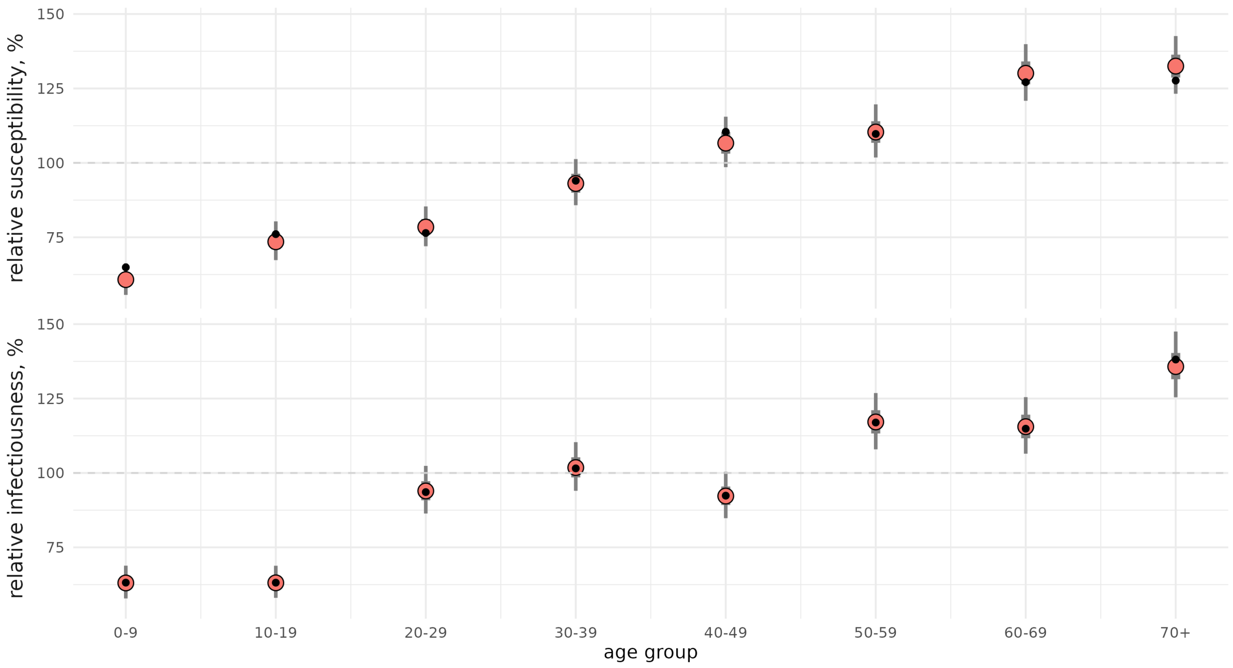


Figure 4: Estimation of age-specific susceptibility and infectiousness from simulated data. Black dots indicate the true values used in the simulation (‘ground truth’).

Table 2: True (input) and estimated values for susceptibility and infectiousness for each age group normalized by the population average in the simulation study, as well as the percentage difference between them.

| type | age group | true value | estimated value | difference (%) |
|---|---|---|---|---|
| inf | 0-9 | 0.632 | 0.630 | 0.18 |
| inf | 10-19 | 0.631 | 0.631 | 0.09 |
| inf | 20-29 | 0.936 | 0.939 | 0.40 |
| inf | 30-39 | 1.015 | 1.018 | 0.26 |
| inf | 40-49 | 0.924 | 0.922 | 0.14 |
| inf | 50-59 | 1.170 | 1.171 | 0.15 |
| inf | 60-69 | 1.149 | 1.155 | 0.59 |
| inf | 70+ | 1.381 | 1.357 | 1.71 |
| sus | 0-9 | 0.649 | 0.608 | 6.37 |
| sus | 10-19 | 0.761 | 0.735 | 3.38 |
| sus | 20-29 | 0.764 | 0.784 | 2.61 |
| sus | 30-39 | 0.940 | 0.931 | 1.00 |
| sus | 40-49 | 1.105 | 1.066 | 3.46 |
| sus | 50-59 | 1.097 | 1.104 | 0.58 |
| sus | 60-69 | 1.271 | 1.301 | 2.33 |
| sus | 70+ | 1.277 | 1.325 | 3.80 |

### 4 Ascertainment rates and asymptomatic infections

Not all cases and transmission pairs are detected (ascertained) in surveillance data, and asymptomatically infected individuals may be less infectious than symptomatically infected individuals [6]. In this section we show that if ascertainment is dependent on the age group only, then the estimates of relative susceptibility absorb ascertainment rates, while the estimates for relative infectiousness will still reflect the biological relative infectiousness. Likewise, we show that asymptomatic infections cannot be disentangled from the estimates of susceptibility, but do not affect infectiousness.

Let $r_i$ denote the ascertainment probability of infected individuals in age group $i$, $i = 1, \dots, m$. When the ascertainment probabilities are only age-group dependent, then an infectee of age group $i$ reports an infector of age group $j$ according to the reporting probability $r_j$ independent of $i$. Therefore, the number of ascertained transmission pairs are

$$\tilde{P}_{ij}^{T} = r_i r_j P_{ij}^{T},$$

and the ascertained infected cases

$$\tilde{Y}_j^* = r_j Y_j^*.$$

Then, the data on ascertained transmission pairs and ascertained cases gives

$$\tilde{P}_{ij} \sim c_{ij} S_i I_j r_i r_j X_i^* Y_j^* = c_{ij} S_i I_j r_i X_i^* \tilde{Y}_j^*.$$

Therefore, the estimators for susceptibility and infectiousness after correcting for age-specific ascertainment rate are

$$r_i S_i = \sum_{j=1}^{m} \frac{\tilde{P}_{ij}}{c_{ij} X_i^* \tilde{Y}_j^*}$$

and, normalizing $\sum_{i=1}^{m} r_i \, S_i = 1$,

$$I_j = \sum_{i=1}^{m} \frac{\tilde{P}_{ij}}{c_{ij} X_i^* \tilde{Y}_j^*}.$$

Therefore, unless additional external estimates for ascsertainment probabilities $r_k$ are available, the estimates for age-specific susceptibility absorb the effects of ascertainment, while the estimates for age-specific infectiousness will still reflect the true underlying biological infectiousness.

If, in addition, we assume that there is an age-dependent probability for an infective individual to be symptomatic ($0 \leq q_i \leq 1$) and that only symptomatic individuals are ascertained, independent of the ascertainment rate, then

$$\tilde{P}_{ij} \sim c_{ij} S_i I_j r_i q_i X_i^* \tilde{Y}_j^*,$$

where the ascertained infected cases are $\tilde{Y}_j^* = r_j q_j Y_j^*$. Again, the estimates for age-specific susceptibility absorbs both reporting probabilities and probabilities to be symptomatic:

$$q_i r_i S_i = \sum_{j=1}^{m} \frac{\tilde{P}_{ij}}{c_{ij} X_i^* \tilde{Y}_j^*},$$

while, normalizing $\sum_{i=1}^{m} q_i \, r_i S_i = 1$, the estimates for age-specific infectiousness remain unchanged:

$$I_j = \sum_{i=1}^{m} \frac{\tilde{P}_{ij}}{c_{ij} X_i^* \tilde{Y}_j^*}.$$

#### 5 Wild type vs Alpha period

In the main analysis we considered the period from 1 July 2020 until 31 March 2021. During this period in The Netherlands, both the wild type of SARS-CoV-2 and the Alpha variant were dominant at different times. In this sensitivity analysis we show that the estimates do not change qualitatively if we split into a wild type and Alpha period instead. Figure S5 compares three sets of estimates: i) the main analysis of all transmission pairs in the entire period 01-07-2020 until 31-03-2021, ii) estimates with transmission pairs when the wild type was dominant (1 July 2020 until 4 February 2021), and iii) estimates with transmission pairs when the Alpha variant was the dominant type (5 February 2021 until 31 March 2021). Each transmission pair was assigned to a period if the infector tested positive after the start date of that period and the infectee tested positive before the end date of the same period. The results show that the estimates are qualitatively similar whether the analysis is performed for the whole period or for each variant-specific period separately. However, for the youngest age group (0-9 years), susceptibility and infectiousness are lower in the wild type period than in the Alpha period.

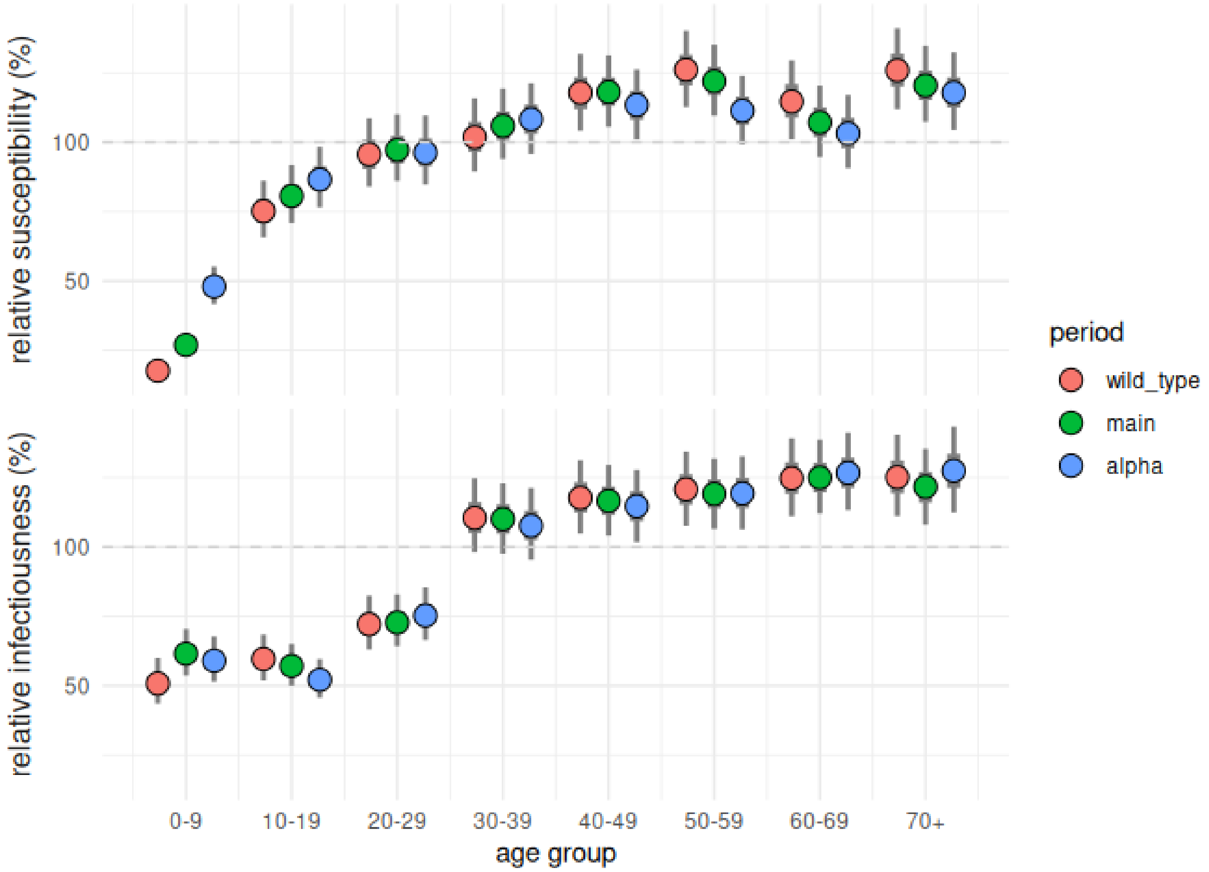


Figure 5: Comparison of age-specific susceptibility and infectiousness estimates normalized to the population average for different time periods: the entire study period as in the 'main' analysis (1 July 2020 - 31 March 2021) and variant-specific periods when the 'wild_type' (1 July 2020 - 4 February 2021) and 'alpha' (5 February 2021 - 31 March 2021) variants were dominant.

#### 6 Impact of age-specific interventions on reducing the reproduction number

We use the next-generation-matrix (NGM) approach [7] to calculate how the reproduction number changes in response to age-specific interventions, both with and without accounting for age-specific susceptibility and infectiousness. These calculations are performed for the initial phase of the epidemic and in absence of any other interventions. Let $\beta$ denote the transmission rate, $\gamma$ the recovery rate, $c_{ij}^{H}$ the household contact rate between an individual of group $i$ and and individual of group $j$, and $c_{ij}^{NH}$ the contact rate for contacts outside of the household. $K$ is the matrix with elements $k_{ij} = (c_{ij}^{H} + c_{ij}^{NH})S_i I_j x_j$ where $x_j$ is the population fraction of group $j$. The NGM for an age-structured SEIR is $(\beta/\gamma)K$, and the reproduction number is the dominant eigenvalue of the NGM $(\beta/\gamma)K$.

When age-specific interventions are applied, contact rates outside the household for the targeted age groups are reduced. The NGM is altered accordingly, and the reproduction number under the intervention can be calculated for the altered NGM. Let $\omega_j$ denote the fraction reduction in contacts outside of the household. Then the altered contact rates become $\tilde{c}_{ij} = c_{ij}^{H} + (1-\omega_i)(1-\omega_j)c_{ij}^{NH}$ and the corresponding NGM $(\beta/\gamma)\widetilde{K}$.

Let $\rho(.)$ denote the dominant eigenvalue of a matrix. The proportional reduction in the reproduction number due to an intervention is $1 - \rho(\widetilde{K})/\rho(K)$. Note that transmission parameters $\beta$ and $\gamma$ are omitted as they cancel out in calculating the proportional reduction. The reduction in the reproduction number is calculated using the age-specific infectiousness and susceptibility as was estimated in the main analysis and using a uniform susceptibility and infectiousness over all age groups.

In the school closure scenario, we assumed that the first two age groups (0–19 years) reduced their out-of-household contacts by 65% [8], $\omega_i = 0.65$ for $i = 1,2$ and $\omega_i = 0$ otherwise. The resulting reduction in the reproduction number for the school-closure scenario was 29% when age differences in susceptibility and infectiousness were ignored, but only 8% when these age differences were included. For the work-from-home measures, we assumed that age groups 3-6 (20–59 years) reduced their out-of-household contacts by 40% [9], reflecting an assumption that about half of their contacts occur at work. Thus $\omega_i = 0.40$ for $i = 3, \dots, 6$ and $\omega_i = 0$ otherwise. In this case, the reduction in the reproduction number was 20% when age differences in susceptibility and infectiousness were ignored, but 41% when they were accounted for.
